# Investigating laboratory capacity and whole genome sequencing practices to inform standardised protocols for bacterial genomic surveillance in Africa

**DOI:** 10.64898/2026.09.24.26363966

**Authors:** Chanté Fortuin, Larisse Bolton, Angela Dramowski, Ebenezer Foster-Nyarko, Kathryn E. Holt, Mae Newton-Foot, Charlene Rodrigues, Andrew Whitelaw

## Abstract

Sepsis is a substantial contributor to neonatal mortality in Africa, with *Klebsiella pneumoniae* as the leading causative pathogen. Incorporating molecular tools such as whole genome sequencing (WGS) into routine surveillance of important bacterial pathogens, including those causing neonatal sepsis, can support targeted deployment of infection prevention and control measures and treatment interventions. This study aimed to understand the current practices and capacity for culture, identification and genomic analysis of bacterial pathogens in Africa, with a particular focus on *K. pneumoniae,* to support standardised genomic surveillance.

A cross-sectional online survey was distributed to various clinician and scientist networks across Africa. All respondents completed a demographics section, while sections on laboratory test capacity, blood culture practices, WGS, and bioinformatics analysis were completed based on experience in these aspects.

Responses were received from 52 institutions in 25 African countries. Among 51 institutions completing the laboratory test capacity section, 61% reported performing blood cultures, with the majority using manual commercial biochemical systems for species identification (71%) and manual disk diffusion for antibiotic susceptibility testing (82%). Participating institutions reported near-universal access to a –80°C freezer, centrifuge and thermocycler, but shaking incubators were less commonly available (67%).

More than half of respondents (54%) indicated that they perform the entire WGS workflow (including DNA extraction, library preparation and WGS) internally, with most performing DNA extraction internally (88%) using column-based kits (70%). Outsourcing was progressively more common at later workflow stages. Infrastructure challenges that limit bioinformatics analysis capabilities were reported by all 23 respondents who completed this question, including limited access to high-performance computing and data storage space. This survey found sufficient capacity to support genomic surveillance of neonatal sepsis pathogens across participating African institutions. However, investments in blood culture, supply chains, sequencing, bioinformatics, and data dissemination capacity are needed to realise its full potential. These findings will support the development of standardised genomic surveillance protocols tailored to the diverse capacities of African laboratories.

## INTRODUCTION

Sepsis remains a leading cause of mortality among neonates (newborns up to 28 days old) in Africa, with a disproportionate burden concentrated in regions where capacity for prevention, diagnosis, and treatment is critically limited (1–4). Across sub-Saharan Africa, neonatal mortality rates continue to exceed the Sustainable Development Goal target of 12 deaths per 1000 live births (5), underscoring the urgent need for targeted, evidence-based interventions tailored to resource-constrained healthcare settings.

Globally, Gram-negative pathogens are the leading cause of both early– and late-onset neonatal sepsis, with *Klebsiella pneumoniae* being the single most common causative pathogen (6, 7), associated with high rates of antimicrobial resistance (AMR) (8). *K. pneumoniae* is therefore a high-priority target for genomic surveillance using whole genome sequencing (WGS), which can enable sub-species-level characterisation of serotypes, virulence factors, AMR determinants, and clonal lineages. This is essential to track the transmission of drug-resistant clones across geographical settings, guide infection prevention and control (IPC) measures and public health responses, and inform targeted interventions such as treatments and vaccine development (9, 10).

Comprehensive surveillance of bacterial pathogens requires a multi-level approach, integrating WGS with traditional microbiological culture and antimicrobial susceptibility testing (AST). Standardisation of routine blood culture, species identification and AST methods and data reporting is essential for effective surveillance of pathogen distribution and AMR trends across settings (11, 12). Likewise, standardised WGS and bioinformatics analysis workflows are similarly important to ensure consistent generation and interpretation of genomic data. DNA extraction from encapsulated Gram-negative bacilli may be challenging due to capsular exopolysaccharides (13). The hypermucoviscous phenotype of some *K. pneumoniae* strains present further challenges for DNA extraction by complicating sedimentation during centrifugation, as well as cell lysis (14, 15). Therefore, protocols adapted to mitigate the challenges presented by mucoid organisms such as *K. pneumoniae* are needed to support genomic surveillance of this organism.

In this study, we aimed to understand the current practices and capacity for blood culture, identification and genomic analysis of bacterial pathogens in Africa, with a particular focus on *K. pneumoniae*. Through this analysis, we aim to inform development of standardised protocols tailored to the capacity of African institutions, to support sustainable genomic surveillance of important bacterial pathogens, including those causing neonatal sepsis.

## METHODS

### Study context

The KlebNET Genomic Surveillance Platform (KlebNET-GSP) (16), established in 2021, aims to integrates key informatics tools, resources and data to help build genomic surveillance capacity for K. pneumoniae, with the aim of supporting vaccine development to prevent neonatal sepsis (17). NeoNET AFRICA is a multidisciplinary network of clinicians, researchers and public health specialists focussing on optimising neonatal sepsis prevention, diagnosis and treatment (18, 19). KlebNET-GSP has partnered with NeoNET AFRICA to coordinate development of genomic surveillance capacity for neonatal sepsis pathogens, including the development of protocols with Klebsiella-specific considerations where needed. Although the activities of these networks have a specific focus on neonatal sepsis and K. pneumoniae, the survey investigated the broader laboratory and genomic surveillance capacity applicable to bacterial pathogens in general.

### Study design

A cross-sectional online survey was created in REDCap (20, 21), aimed at clinicians and scientists from across Africa involved in microbiological laboratory testing and/or WGS of neonatal sepsis-causing bacterial pathogens. The survey comprised the following key sections: 1) Demographics, 2) Laboratory test capacity, 3) Practices for genomic sequencing of bacterial pathogens, and 4) Practices for bioinformatics analysis of bacterial whole genome sequences. All participants were required to complete the demographics section, while the completion of subsequent sections was determined by filtering questions based on experience.

The survey questions were a combination of multiple-choice, often allowing the participant to provide more than one answer, and dichotomous “yes” or “no”. Free-text answers were only required where additional information was needed. As far as possible, questions were set to require a response to minimise missing data. The survey and accompanying information were available in English (Appendix S1), French and Portuguese.

Participant recruitment followed an initial purposive sampling approach, with survey dissemination focused on existing neonatal sepsis clinicians and scientist networks across Africa such as NeoNET AFRICA (18, 19), the Infection Control Africa Network (ICAN) (22), the African Society for Laboratory Medicine (ASLM) (23), Pasteur Africa Network (24) and H3Abionet (25, 26). The survey was also advertised across various social media and academic platforms. Thereafter, participant recruitment followed a snowball sampling approach through referrals from respective participants. The survey remained active for 3 months, from 8 August 2025 until 10 November 2025.

### Ethical considerations

Ethical approval for the study was obtained from the Stellenbosch University Health Research Ethics Committee (HREC; Ref no: N25/04/040). All participants were required to provide written informed consent prior to completion of the survey. Survey responses were voluntary and anonymous, unless respondents agreed to provide their contact information for further involvement.

### Data analysis

The survey responses were analysed using Microsoft Excel, as well as RStudio (version 2026.01.1+403) running R statistical software version 4.5.2 (27). All responses were categorical in nature and thus analysed using frequency measures such as counts or percentages. Where necessary, results were stratified according to type of laboratory. Countries were grouped by African region according to the United Nations classification (28). When different sections of the survey were completed by individuals from the same institution, the sections were combined into one response. Where two individuals from the same institution completed the same section of the survey, answers were combined for questions with multiple answers allowed and the greater number category was used for questions allowing a single answer.

## RESULTS

### Demographics

A total of 58 survey responses were received, all of which included a completed demographics section. A third of respondents (32.8%; n = 19/58) indicated their job role as research scientist (Fig A in S2 Appendix). A total of 52 unique institutions were represented across 25 African countries, predominantly in Eastern and Western Africa (Fig. 1). South Africa (13.5%; n = 7/52), Ghana (11.5%; n = 6/52) and Zambia (9.6%; n = 5/52) accounted for the highest proportion of responses. Respondents were able to select more than one laboratory type, with research laboratory (57.7%; n = 30/52) being the most common, followed by clinical diagnostic laboratory (36.5%; n = 19/52) and national public health laboratory (26.9%; n = 14/52).

**Fig. 1:**
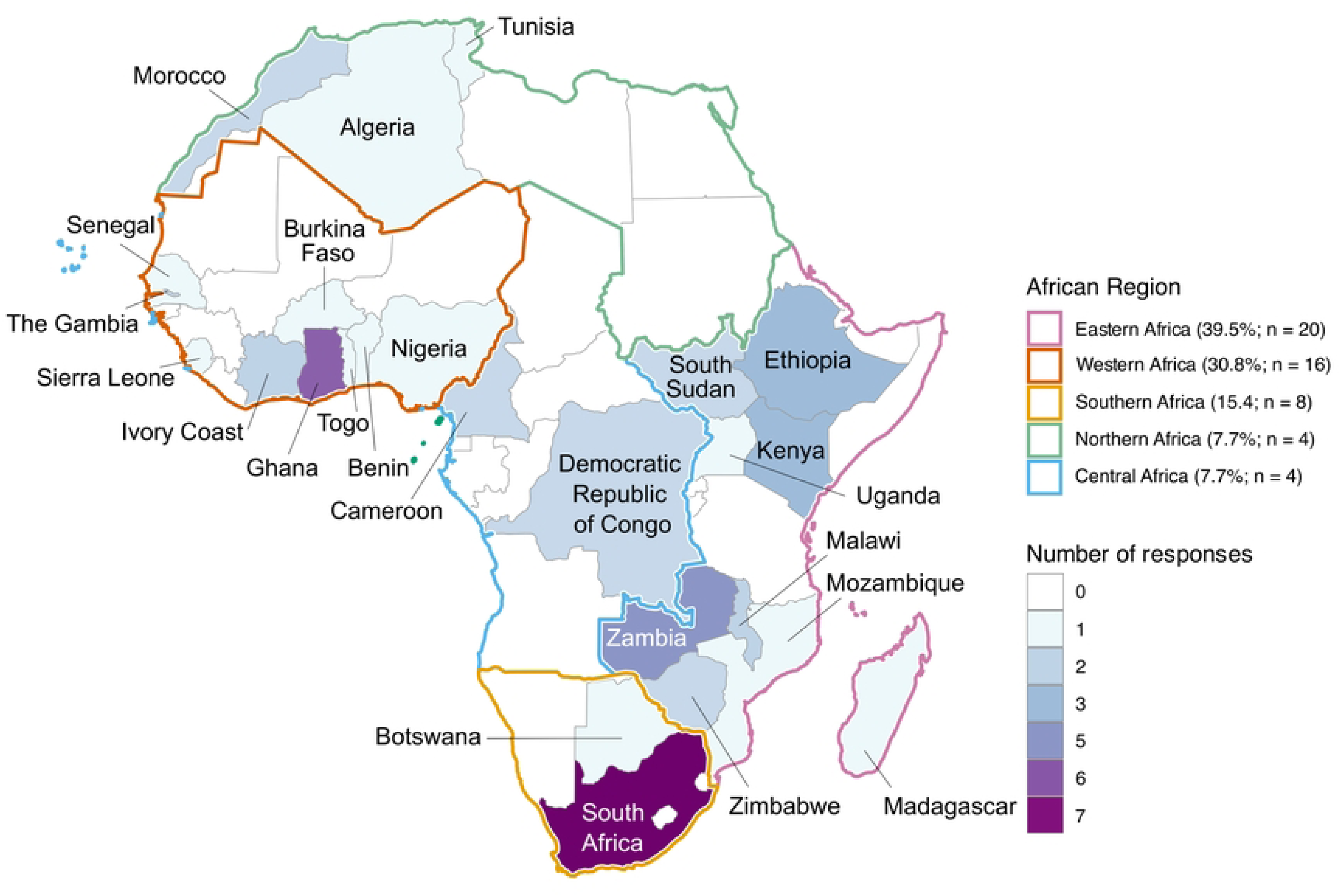
Country distribution of the 52 institutions across Africa that provided responses to the survey, stratified by UN-defined African Region.

### Laboratory test capacity

The laboratory test capacity section was completed by respondents from all but one of the 52 institutions (98.1%; n = 51). Of these, 61.5% (n = 32/51) indicated that their laboratory performs blood cultures and completed a subsection of questions on blood culture practices. Of institutions performing blood cultures, more than half (56.2%; n = 18/32) were clinical diagnostic laboratories, followed by research laboratories (43.7%; n = 14/32) and national public health laboratories (31.2%; n = 10/32), including respondents selecting each laboratory type alone or in combination with other laboratory types. Manual blood culture methods (43.8%; n = 14/32) and automated systems (40.6%; n = 13/32) were both common, with 15.6% (n = 5/32) of respondents indicating the use of both methods. Commercial blood culture media (62.5%; n = 20/32) was more frequently used than in-house prepared media (25.0%; n = 8/32), with the use of both media types indicated by 12.5% (n = 4/32) of respondents.

Most respondents indicated that their laboratories process 11-50 blood cultures per month (37.5%; n = 12/32). This option accounted for the largest proportion of responses from all laboratory types: research laboratories (50.0%; n = 7/14), clinical diagnostic laboratories (38.9%; n = 7/18) and national public health laboratories (30%; n = 3/10) (Fig B in S2 Appendix). The greatest proportion of respondents indicated that 6-10 blood cultures test positive per month (28.1%; n = 9/32). The combination of responses for the number of blood cultures tested and positive are given in Fig.2a, with 11-50 processed per month and yielding 6-10 positives being the most common pattern. When asked which pathogens were frequently (at least once per month) isolated from blood cultures, most respondents selected *Staphylococcus aureus* (96.9%; n = 31), followed by *Escherichia coli* (78.1%; n = 25) and *Klebsiella* spp. (75.0%; n = 24) (Fig. 2b).

**Fig. 2:**
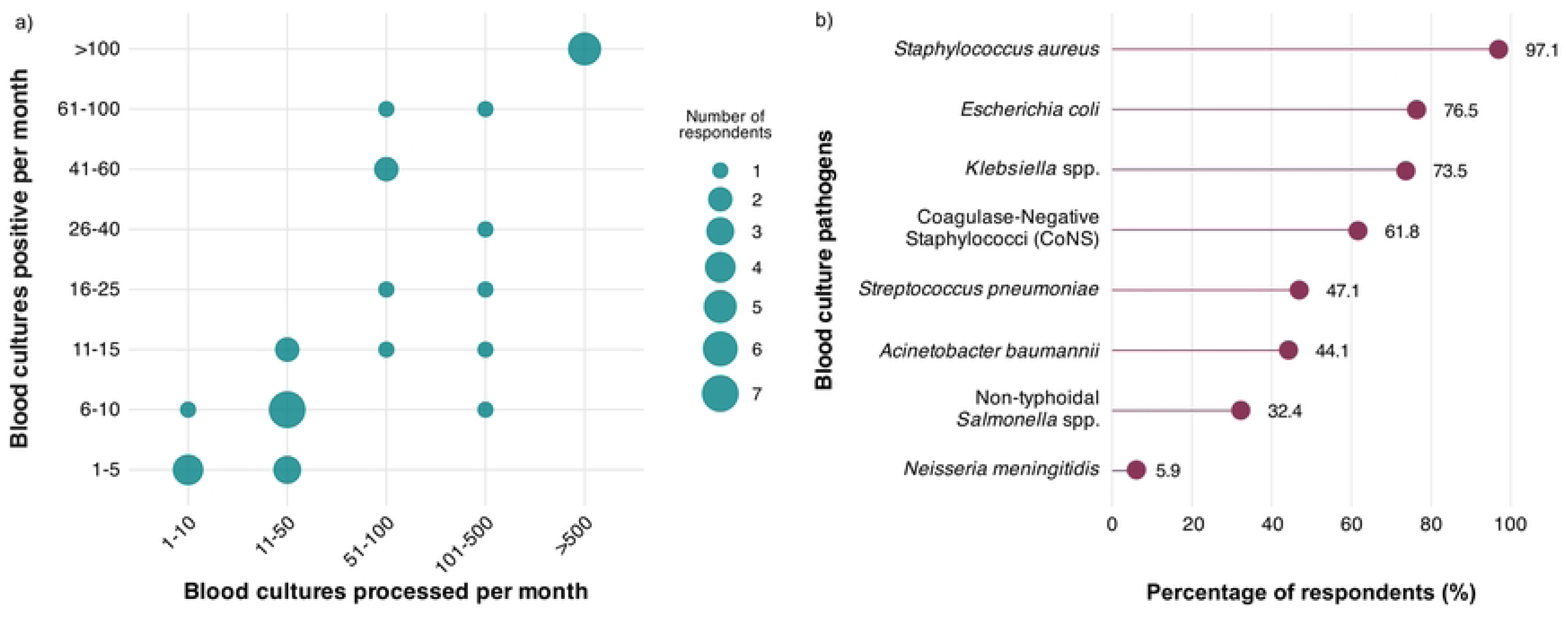
Blood culture practices across 32 institutions that completed the blood culture practices section. (a) Number of respondents indicating each combination of number of blood cultures processed per month and number of positive blood cultures per month. (b) Species indicated as being frequently (at least once per month) isolated from blood cultures, with multiple answers allowed.

Questions on species identification and AST practices were completed by all 51 respondents in the laboratory capacity section. Overall, the most commonly used methods for species identification were manual commercial biochemical systems (70.6%; n = 36/51), followed by PCR or other nucleic acid-based methods (68.6%; n = 35/51). Automated biochemical identification systems and serotyping were equally prevalent (both 41.2%; n = 21/51), while Matrix-Assisted Laser Desorption/Ionization Time-of-Flight Mass Spectrometry (MALDI-TOF MS) was the least frequently used (21.6%; n = 11/51).

Notably, species identification methods differed by laboratory type (Fig.3a), with national public health laboratories reporting more frequent use of automated biochemical identification systems (78.6%; n = 11/14) and serotyping (71.4%; n = 10/14) compared to other laboratory types. The use of PCR or other nucleic acid-based methods was less common in clinical diagnostic laboratories, which mostly reported the use of manual commercial biochemical systems (78.9%; n = 15/19). MALDI-TOF MS was the least commonly used identification method across all laboratory types (<25%).

For AST, the most common method reported overall was manual disk diffusion (82.4%; n = 42/51), followed by automated systems (39.2%; n = 20/51), gradient diffusion (35.3%; n = 18/51), and broth microdilution / agar dilution (25.5%; n = 13/51). The use of AST methods was similar across all laboratory types, although automated systems were less common in research laboratories compared to other laboratory types (Fig.3b). Among respondents selecting “Other” AST methods, two research laboratories and one clinical diagnostic laboratory indicated that they do not perform AST, while two national public health laboratories and one research laboratory indicated that they use genomics to infer AST. One respondent who selected more than one laboratory type (clinical diagnostic and research laboratory) indicated that they outsource AST.

### WGS practices

Of the 52 institutions, 41 (78.8%) indicated that their laboratory performs WGS; however, only 33 respondents were able to complete the WGS practices section (63.5% of all 52 institutions; 80.5% of the 41 that perform WGS). When asked how many bacterial genomes their laboratory had sequenced in the last five years, a large proportion of respondents indicated greater than 500 (36.4%; n = 12/33) or between 101 and 500 (21.2%; n = 7/33). Respondents from research laboratories (39.1%; n = 9/23) and national public health laboratories (40.0%; n = 4/10) mostly indicated greater than 500 genomes, while majority of those from clinical diagnostic laboratories indicated between 101 and 500 (50.0%; n = 5/10) (Fig C in S2 Appendix).

Respondents were asked whether they perform DNA extraction, library preparation and WGS internally or externally and were allowed to select both of these options. More than half of respondents (54.5%; n = 18/33) indicated that they perform all three steps internally only, with majority performing DNA extraction internally only (87.9%; n = 29/33) and the amount of outsourcing increasing as these steps progress (Fig. 4).

**Fig. 3:**
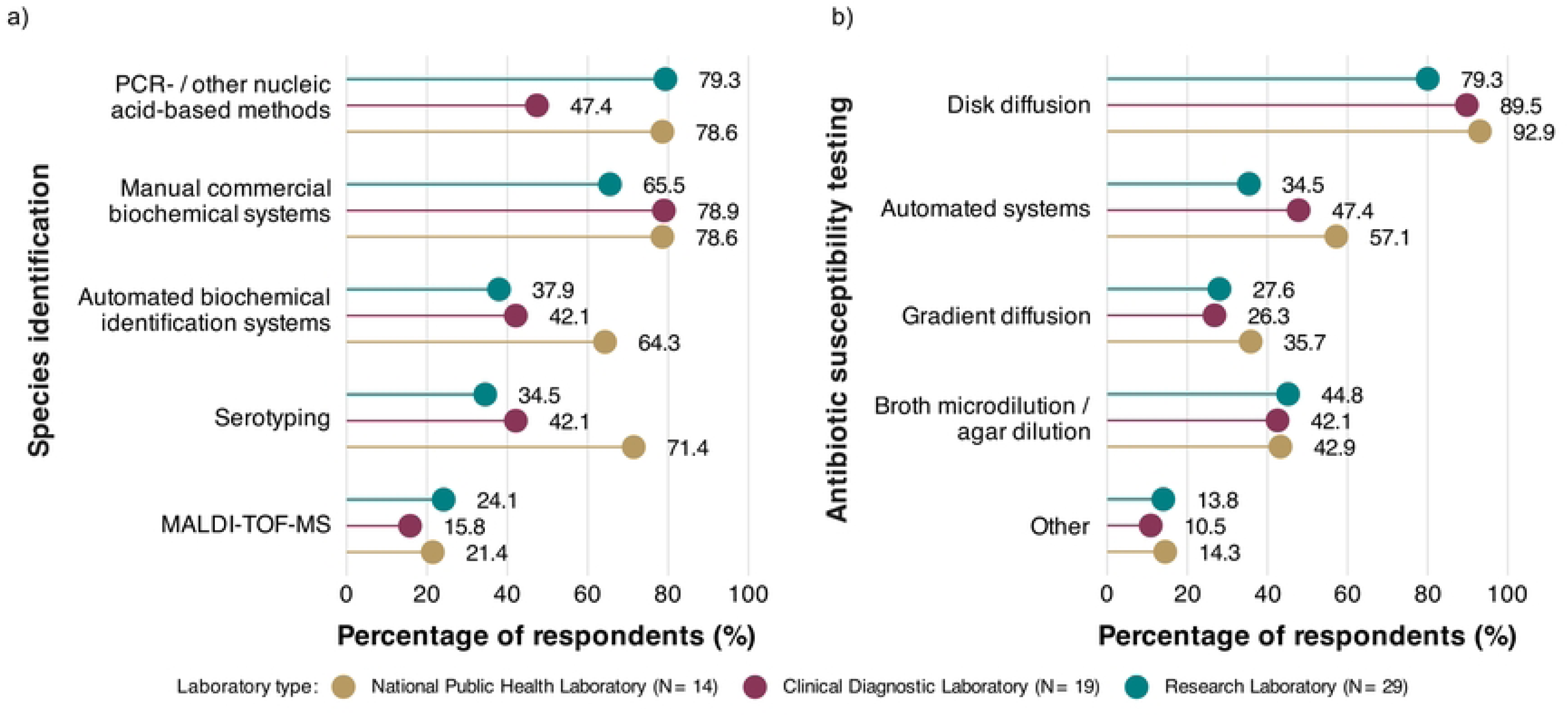
Species identification (a) and antibiotic susceptibility testing (b) methods performed across 51 institutions that completed the laboratory capacity section, stratified by laboratory type. Other = includes responses such as “None”, “Genomics” and “Outsourced”.

**Fig. 4:**
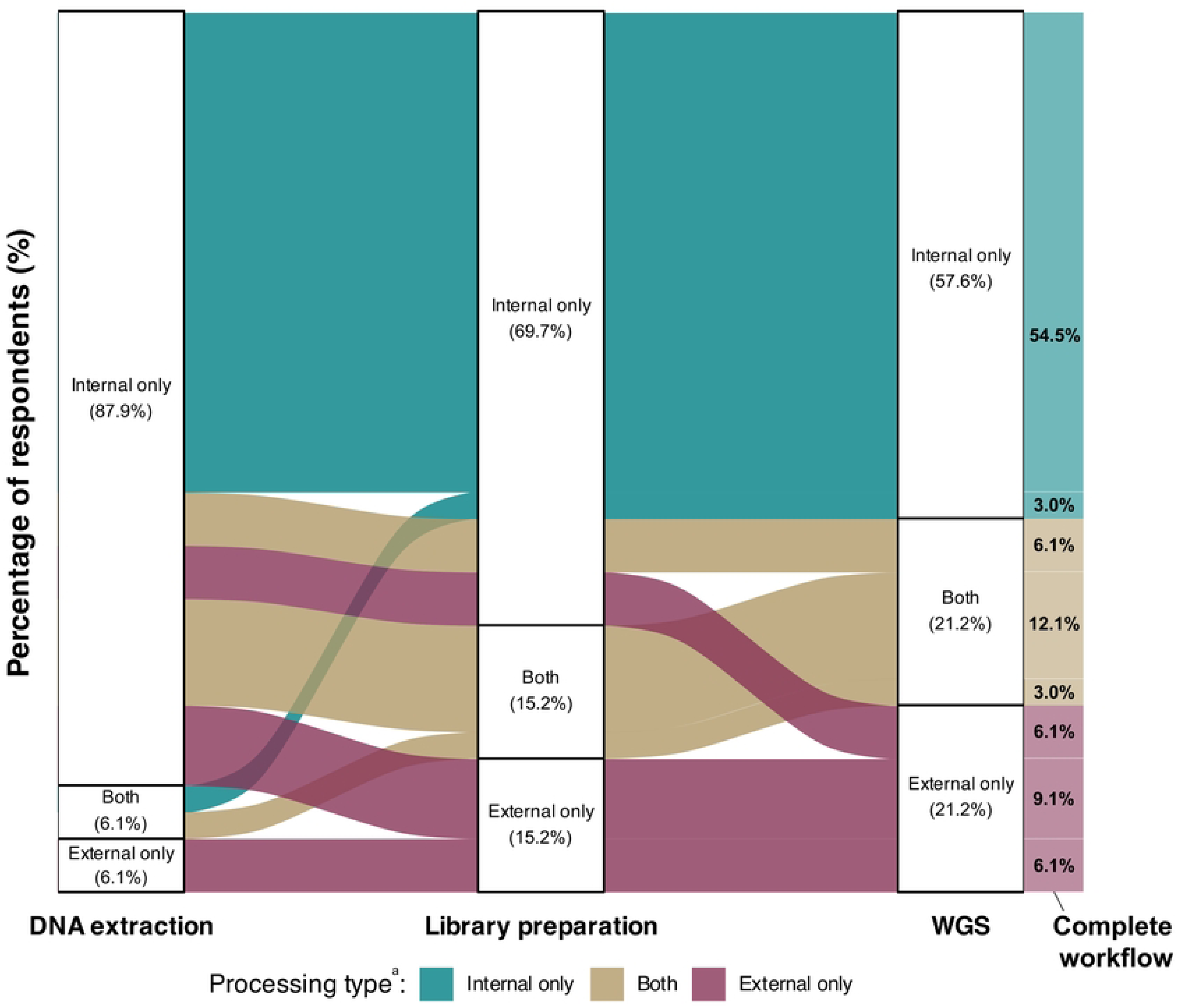
Workflows for DNA extraction, library preparation, and whole genome sequencing (WGS) reported by 33 respondents who completed the WGS practices section. Each column represents a processing step, with strata indicating the percentage of respondents performing each step only internally, only externally, or using both approaches. The widths of the flows represent the number of respondents following each complete workflow pathway, with percentages indicated on the far right. ^a^ Flows are coloured according to processing type in the WGS column.

In terms of equipment access, all 33 respondents indicated that they have access to a centrifuge in their laboratory and most have access to a –80°C freezer and thermocycler, with only one clinical diagnostic laboratory not having access to the latter two items (Fig C in S2 Appendix). A shaking heating block was less commonly available at all three laboratory types (60 – 80%); however, each respondent had access to at least one type of heating instrument, albeit without shaking.

Most respondents indicated the use of column-based kits for DNA extraction, mainly those manufactured by Qiagen, while other manual or automated methods were also common (Table 1). Column-based kits was the most frequently selected method in research laboratories (78.3%; n = 18/23) and clinical diagnostic laboratories (60%; n = 6/10), while automated methods were more common in national public health laboratories (70%; n = 6/10) compared to the other laboratory types (Fig C in S2 Appendix).

**Table 1:**
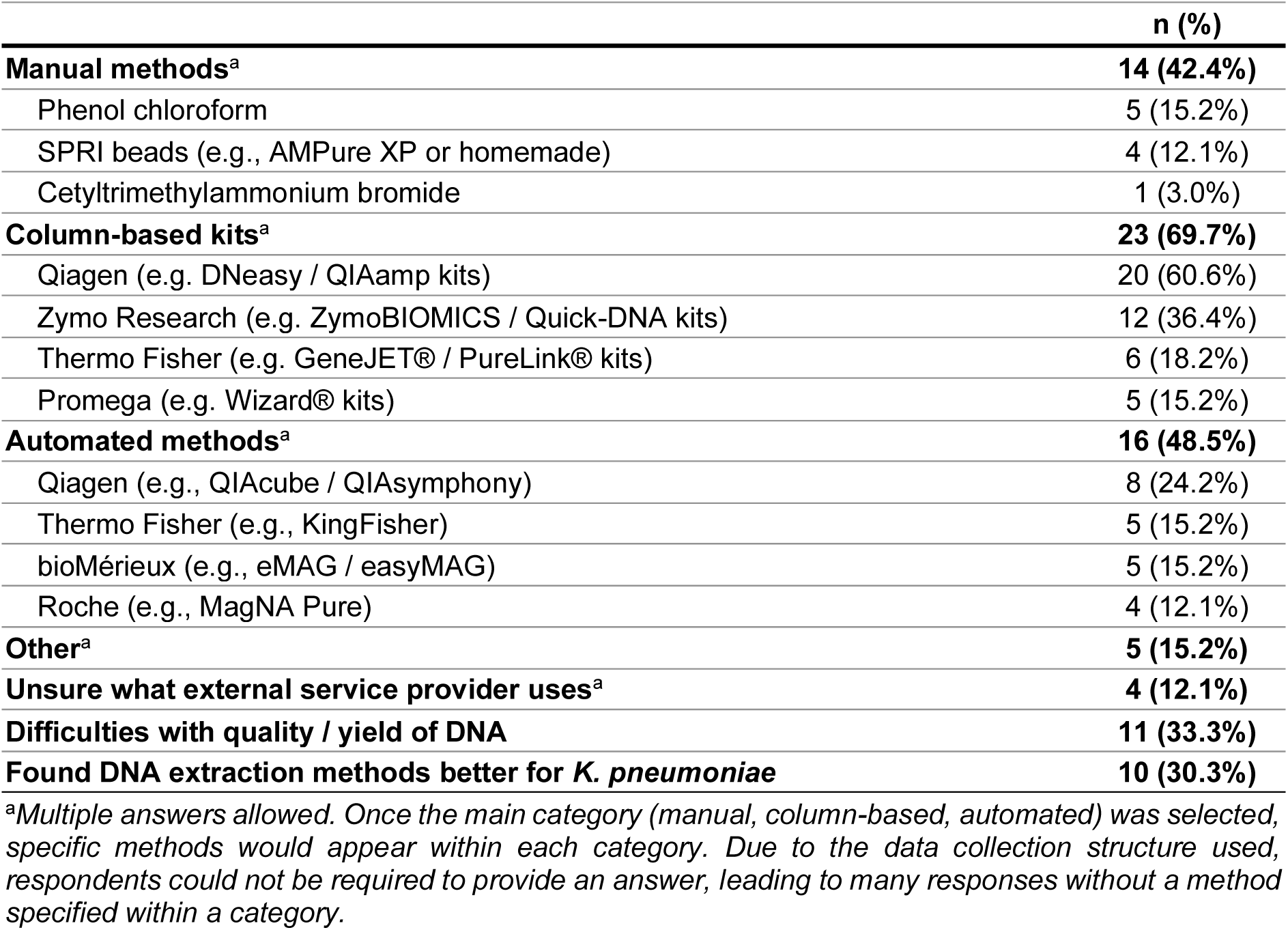
DNA extraction practices of 33 survey respondents completing the WGS practices section.

A third of respondents (33.3%; n = 11/33) reported that they have experienced difficulties with the yield or quality of DNA obtained from certain bacterial pathogens. These challenges were mostly experienced with Gram-positive cocci (n = 4) and *Mycobacterium tuberculosis* (n = 2), with some reporting difficulty with capsule-forming bacteria (n = 1) and hypermucoviscous *Klebsiella* spp. (n = 1). Similarly, 30.3% (n = 10/33) of respondents indicated that they have found certain DNA extraction methods to work better for *K. pneumoniae*, with six of these specifying Qiagen column-based kits.

Illumina and Oxford Nanopore Technologies (ONT) were the most commonly accessed sequencing platforms, either internally or through an external service provider (Table 2). The most frequently used instruments for each of these two platforms, both internally and externally, were the Illumina MiSeq and the ONT MinION. Other platforms were rarely used (Table 2).

**Table 2:**
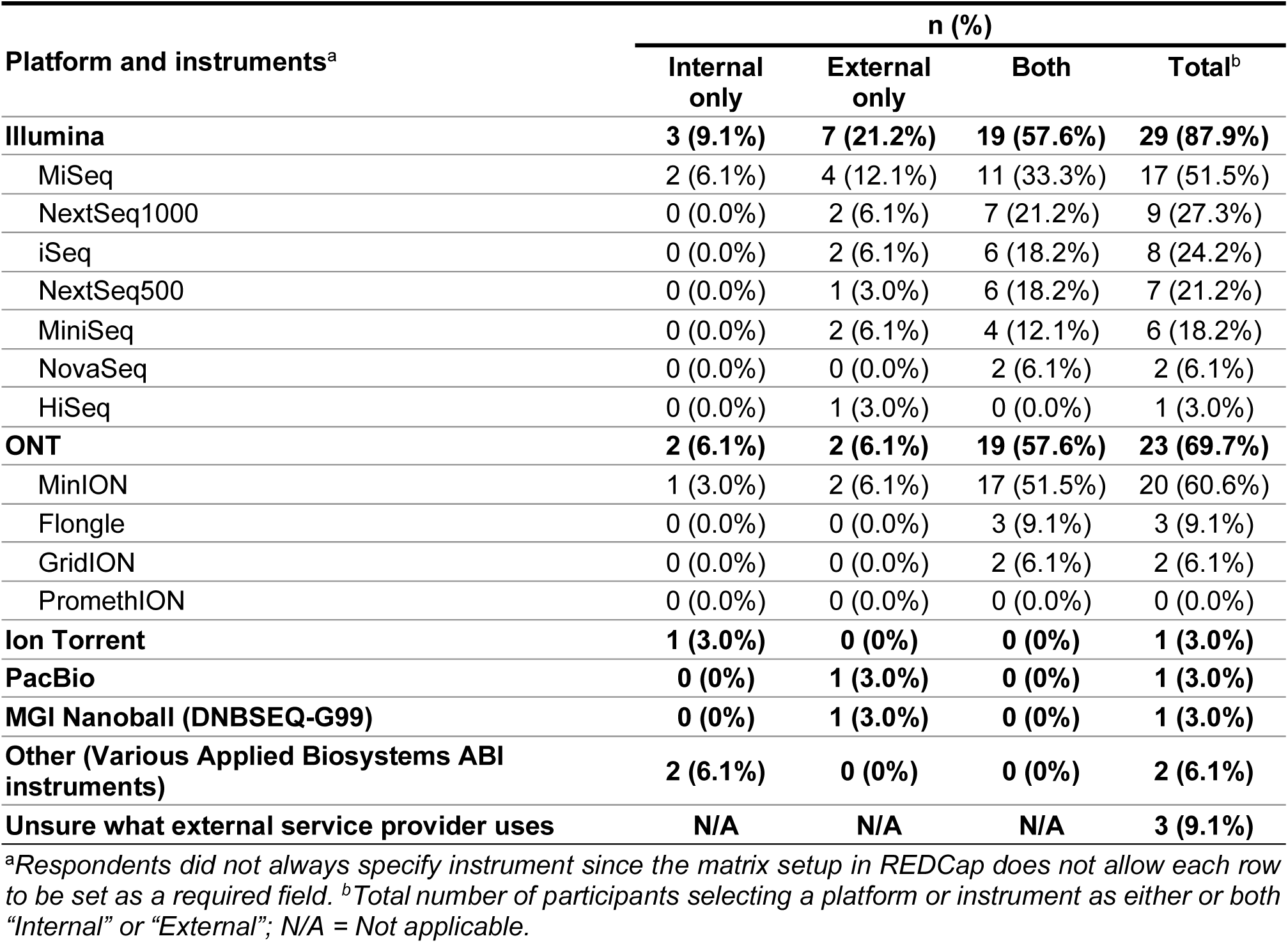
WGS platforms and instruments accessed (internally, externally or both) by 33 survey participants who completed the WGS practices section.

| Platform and instruments<sup>a</sup> | n (%) |  |  |  |
| --- | --- | --- | --- | --- |
|  | Internal only | External only | Both | Total<sup>b</sup> |
| **Illumina** | **3 (9.1%)** | **7 (21.2%)** | **19 (57.6%)** | **29 (87.9%)** |
| MiSeq | 2 (6.1%) | 4 (12.1%) | 11 (33.3%) | 17 (51.5%) |
| NextSeq1000 | 0 (0.0%) | 2 (6.1%) | 7 (21.2%) | 9 (27.3%) |
| iSeq | 0 (0.0%) | 2 (6.1%) | 6 (18.2%) | 8 (24.2%) |
| NextSeq500 | 0 (0.0%) | 1 (3.0%) | 6 (18.2%) | 7 (21.2%) |
| MiniSeq | 0 (0.0%) | 2 (6.1%) | 4 (12.1%) | 6 (18.2%) |
| NovaSeq | 0 (0.0%) | 0 (0.0%) | 2 (6.1%) | 2 (6.1%) |
| HiSeq | 0 (0.0%) | 1 (3.0%) | 0 (0.0%) | 1 (3.0%) |
| **ONT** | **2 (6.1%)** | **2 (6.1%)** | **19 (57.6%)** | **23 (69.7%)** |
| MinION | 1 (3.0%) | 2 (6.1%) | 17 (51.5%) | 20 (60.6%) |
| Flongle | 0 (0.0%) | 0 (0.0%) | 3 (9.1%) | 3 (9.1%) |
| GridION | 0 (0.0%) | 0 (0.0%) | 2 (6.1%) | 2 (6.1%) |
| PromethION | 0 (0.0%) | 0 (0.0%) | 0 (0.0%) | 0 (0.0%) |
| **Ion Torrent** | **1 (3.0%)** | **0 (0%)** | **0 (0%)** | **1 (3.0%)** |
| **PacBio** | **0 (0%)** | **1 (3.0%)** | **0 (0%)** | **1 (3.0%)** |
| **MGI Nanoball (DNBSEQ-G99)** | **0 (0%)** | **1 (3.0%)** | **0 (0%)** | **1 (3.0%)** |
| **Other (Various Applied Biosystems ABI instruments)** | **2 (6.1%)** | **0 (0%)** | **0 (0%)** | **2 (6.1%)** |
| **Unsure what external service provider uses** | **N/A** | **N/A** | **N/A** | **3 (9.1%)** |

<sup>a</sup>Respondents did not always specify instrument since the matrix setup in REDCap does not allow each row to be set as a required field. <sup>b</sup>Total number of participants selecting a platform or instrument as either or both "Internal" or "External"; N/A = Not applicable.

The most commonly used library preparation method for Illumina sequencing was Nextera kits, followed by Amplicon-based methods (Table 3). For ONT, the Rapid Barcoding kit was the most common, followed by the Native Barcoding and Ligation Sequencing kits. Four respondents indicated that they have found certain library preparation methods to work better for *K. pneumoniae*, with three indicating the Illumina DNA prep kit (formerly known as Nextera DNA Flex) and one indicating the ONT Rapid Barcoding Kit.

**Table 3:** Library preparation methods used by 33 survey respondents who completed the WGS practices section.

|  | n | % of total responses | % of respondents using specific platform |
| --- | --- | --- | --- |
| **Illumina** |  | **N = 33** | **N = 29** |
| Nextera kits (e.g. XT DNA, DNA Flex) | 18 | 54.5% | 62.1% |
| Amplicon-based / PCR-based methods (e.g., 16S, resistance genes, targeted sequencing) | 10 | 30.3% | 34.5% |
| TruSeq kits (e.g. PCR-Free, Nano DNA) | 5 | 15.2% | 17.2% |
| Qiagen QIAseq | 5 | 15.2% | 17.2% |
| NEB kits (e.g. NEBNext Ultra) | 4 | 12.1% | 13.8% |
| Unsure what external service provider uses | 3 | 9.1% | 10.3% |
| **ONT** |  | **N = 33** | **N = 23** |
| Rapid Barcoding kit | 16 | 48.5% | 69.6% |
| Native Barcoding kit | 13 | 39.4% | 56.5% |
| Ligation Sequencing kit | 11 | 33.3% | 47.8% |
| Rapid Sequencing kit | 4 | 12.1% | 17.4% |
| NEBNext Companion Module for ONT Ligation Sequencing | 6 | 18.2% | 26.1% |
| Amplicon-based / PCR-based methods | 7 | 21.2% | 30.4% |
| Ultra-Long DNA Sequencing kit | 1 | 3.0% | 4.3% |
| **Ion Torrent:** Amplicon-based / PCR-based methods | **1** | **3.0%** | **N/A** |
| **PacBio:** Unsure what external service provider uses | **1** | **3.0%** | **N/A** |
| **MGI:** MGIEasy kits (e.g. FS, PCR-free, Universal) | **1** | **3.0%** | **N/A** |

N/A = not applicable.

### Reagent logistics

Both the laboratory capacity and WGS practices sections of the survey contained questions regarding reagent logistics. Most respondents indicated that they wait more than three months for both general laboratory reagents (33.3%; n = 17/51) and WGS reagents (45.5%; n = 15/33) to arrive once ordered (Fig. 5a). A reagent shelf life of six months to a year was most common for general laboratory reagents (58.8%; n = 30/51), while two to six months and six months to a year were equally common for WGS reagents (both 42.4%; n = 14/33; Fig. 5b). When asked to estimate what proportion of their orders experience delayed delivery, for example due to an item being out of stock, shipping delays or customs clearance delays, common responses ranged from less than 10% of orders up to 50% of orders (Fig. 5c). Only one respondent each indicated that they never experience delays with delivery of general laboratory reagents or WGS reagents.

**Fig. 5:**
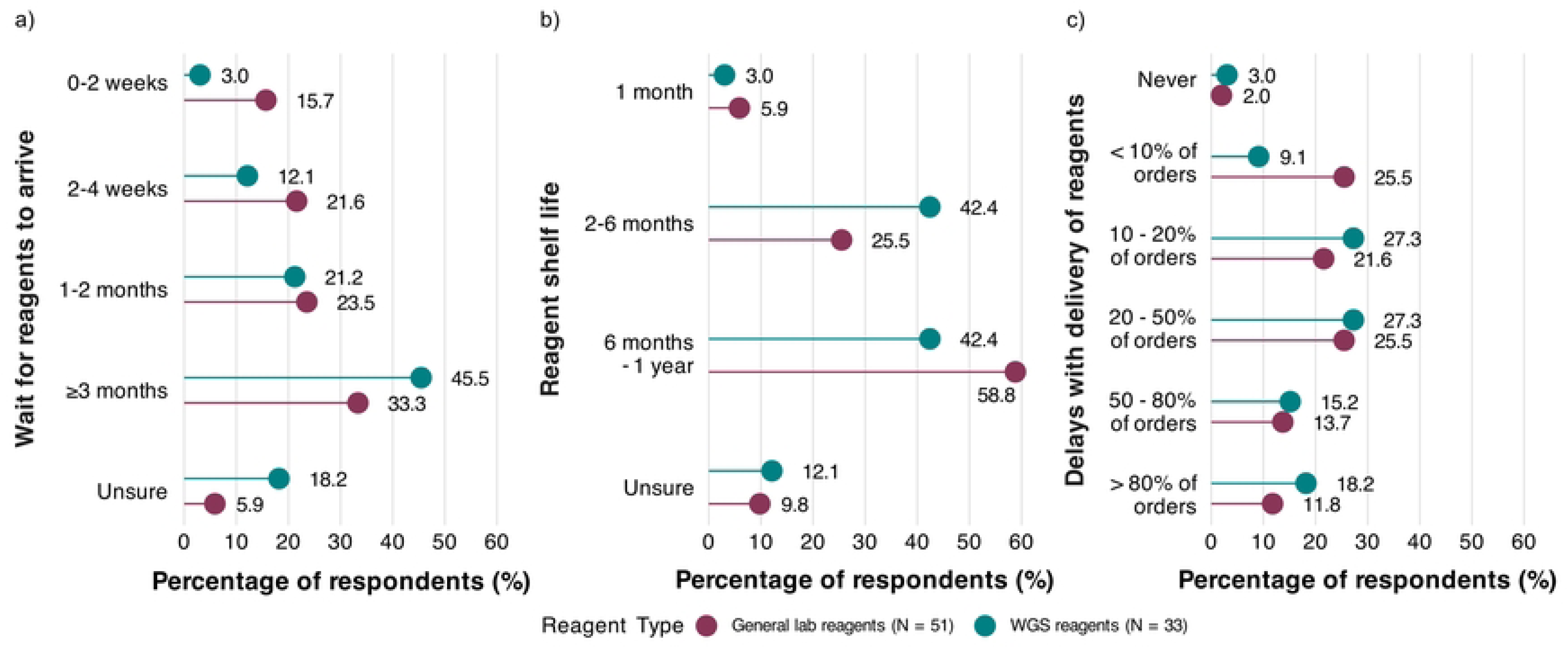
Reagent logistics for general laboratory reagents and WGS reagents. Percentage of respondents indicating (a) how long they wait for reagents to arrive, (b) what the average shelf life is of the reagents and (c) how often they experience delays with delivery of reagents.

### Bioinformatics analysis practises

The bioinformatics analysis section was completed by respondents from 24 institutions (46.1% of the total 52 institutions; 58.5% of the 41 performing WGS). Four of these respondents (16.7%;) indicated that they have experienced difficulties with the quality or yield of sequencing reads, two of whom specified that these were due to DNA extraction or library preparation related issues.

The three most commonly used tools for each step of the analysis process are given in Fig. 6 (see Table A in S2 Appendix for full list). Most respondents (91.7%; n = 22/24) indicated that they use automated workflows such as Galaxy, either internally or through an external service provider. SPAdes, Burrows-Wheeler Aligner (BWA) and Prokka were the most common tools used for assembly, mapping and annotation, respectively. Centre for Genomic Epidemiology (CGE)-based tools (ResFinder, VirulenceFinder, PlasmidFinder) were the most frequently used across various applications such as resistance gene detection, virulence profiling and plasmid analysis (Fig. 6).

**Fig. 6:**
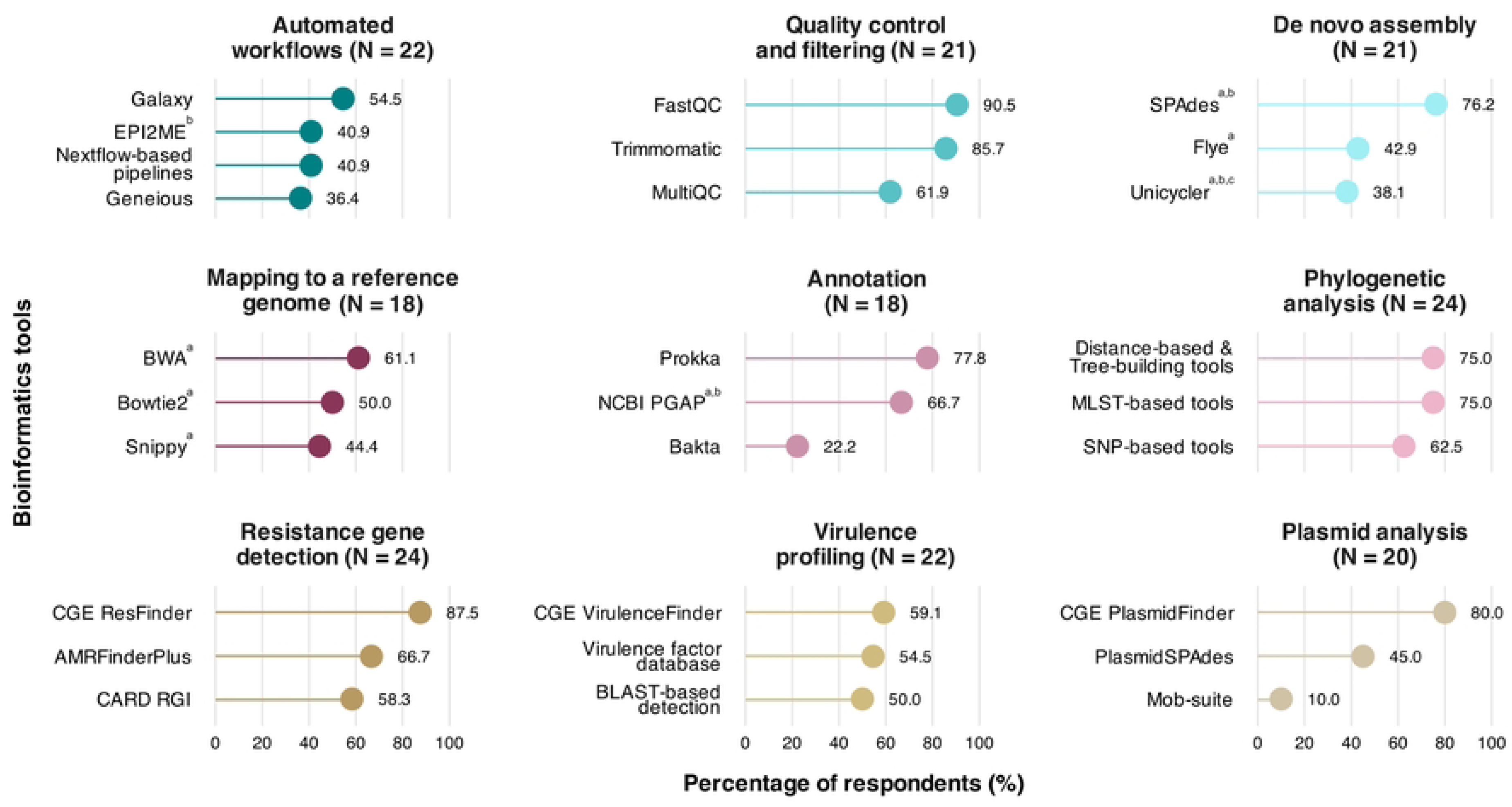
The three most commonly used tools for various bioinformatics analysis processes among 24 survey respondents who completed the bioinformatics practices section. The denominator for the tools within each category is the number of respondents who indicated that they perform that step, as given in the category heading. Multiple answers were allowed. Four tools are listed for Automated workflows since there was a tie for second place. BWA = Burrows-Wheeler Aligner, CGE = Center for Genomic Epidemiology, CARD = Comprehensive Antibiotic Resistance Database, MLST = Multilocus sequence typing, NCBI = National Center for Biotechnology Information, PGAP = Prokaryotic Genome Annotation Pipeline, RGI = Resistance Gene Identifier, SNP = single nucleotide polymorphism. ^a^ Short reads, ^b^ Long reads, ^c^ Hybrid assembly.

Half of the respondents indicated that they make use of in-house servers (50.0%; n = 12/24) to perform bioinformatics analysis, while shared resources (internal or external) were also selected often (both 33.3%; n = 8/24) (Fig. D in S2 Appendix). External hard drives (66.7%; n = 16/24) and computer hard disks (54.2%; n = 13/24) were the most common data storage methods (Fig. D in S2 Appendix). Majority of respondents submit their sequences to one or more public repositories (62.5%; n = 15/24), most commonly the National Center for Biotechnology Information (NCBI; 58.3%; n = 14/24) and the European Nucleotide Archive (ENA; 37.5%; n = 9/24) (Fig. D in S2 Appendix). However, a substantial proportion of respondents (37.5%) reported that they do not submit their sequence data to any public repositories.

Twenty-three respondents provided complete responses to a question on infrastructure challenges that limit their bioinformatics analysis capabilities and all of them indicated one or more challenges. Limited access to high-performance computing (82.6%; n = 19/23) and data storage space (78.3%; n = 18/23) were the most frequently reported, while unreliable internet (60.9%; n = 14/23) and inconsistent electricity (56.5%; n = 13/23) were also common (Fig. D in S2 Appendix).

### Need for increased capacity

All 52 institutions completed a question on their need for increased capacity in certain areas of WGS. Overall, the greatest need was for bioinformatics analysis of WGS data (81.0%; n = 42/52), followed by the actual WGS experiment (71.2%; n = 37/52) and DNA extraction and library preparation (65.0%; n = 34/52). A small proportion of respondents indicated no need for increased capacity (13.0%; n = 7/52), particularly amongst research laboratories and national public health laboratories (Fig. E in S2 Appendix). Clinical diagnostic laboratories indicated similar needs for increased capacity across all three areas.

## DISCUSSION

This study provides a snapshot of the laboratory capacity and WGS practices in African institutions. The survey involved a total of 52 institutions across 25 African countries, mostly in the Eastern and Western African regions. Overall, we found that capacity for genomic surveillance of sepsis pathogens was present across participating African institutions, although reliance on external service providers increased at later stages of the WGS workflow.

Effective genomic surveillance depends on reliable detection and identification of the pathogens of interest, making microbiology laboratory capacity a crucial foundation for genomic analyses. In the laboratory capacity section, 62% of respondents indicated that their laboratory performs blood cultures, leaving a substantial proportion of participating institutions that do not perform this standard diagnostic test for neonatal sepsis. Furthermore, over one-third of laboratories reported processing only 11–50 blood cultures per month, which could suggest limited utilisation of the service even where it is offered. Although our findings represent only 18 diagnostic laboratories, they are consistent with a larger study of 61 hospitals across four African countries (29). That study also found that 62% of hospitals had blood culture capacity, but blood cultures were performed for only 6% of admitted neonates within the three-year study period.

Apart from access challenges, the affordability of blood cultures remains a barrier to increased uptake in most LMICs, especially in sub-Saharan Africa (30–35). As blood cultures are the primary source of bacterial isolates for genomic analyses, limited blood culture capacity may reduce the ability of laboratories to contribute to surveillance programmes. However, the value of blood cultures for patient management as well as surveillance relies not only on access to this test but also on high-quality and comparable microbiological data through standardised specimen collection and laboratory procedures, quality assurance, and reporting practices (29).

Accurate pathogen identification and AST provide the microbiological foundation for genomic surveillance, facilitating appropriate selection of isolates for WGS and enabling comparison of phenotypic and genotypic AMR. Although access to automated pathogen identification and AST platforms such as MALDI-TOF MS and VITEK-II or Microscan was limited among the participating institutions, especially among research laboratories, most respondents reported access to reputable methods such as nucleic acid-based or commercial biochemical identification systems and disk diffusion for AST.

Once a pathogen is isolated in the microbiology laboratory, successful implementation of genomic surveillance also relies on high-quality DNA as input material. DNA extraction does not appear to be a major limiting factor for genomic surveillance capacity among participating laboratories, as evidenced by the fact that 88% of respondents performed DNA extraction in-house, most commonly using column-based extraction kits (70%). These methods are known to produce high-quality DNA suitable for WGS. Furthermore, all respondents in the WGS section reported access to a centrifuge, the primary laboratory equipment required for this method.

However, 65% of all 52 survey respondents indicated a need for increased capacity for DNA extraction and library preparation. The survey combined these two steps as a single option; therefore, the result may reflect a need for increased library preparation rather than DNA extraction capacity, for which respondents reported outsourcing less often than library preparation. Where respondents indicated both internal and external library preparation and WGS, it may relate to different sequencing platforms. For example, they may have access to an ONT platform in-house, which is generally more affordable than Illumina (36), but outsource their Illumina WGS.

Capacity to generate sequencing data is just the first component of genomic surveillance. Translating that data into useful information requires specialised bioinformatics skills and computational resources. The frequent use of CGE-based tools across various applications such as resistance gene detection, virulence profiling and plasmid analysis by 24 survey respondents in the bioinformatics analysis section could reflect the ease of use of the web-based interfaces of these tools compared to others that are mainly implemented through command-line workflows. It may also be a product of training that was provided across various African countries as part of the SeqAfrica project (37), led by the Technical University of Denmark (DTU) where the CGE is based. Substantial investments in genomic pathogen surveillance in recent years, accelerated by the COVID-19 pandemic, has resulted in the launch of many similar projects aimed at expanding sequencing capacity across the African continent (9, 36, 38).

While access to WGS equipment has improved in African countries, barriers to widespread implementation remain, including limited stable internet connectivity, data analysis expertise and storage capacity, as well as their associated costs (9, 36, 38). These challenges were evident in the survey findings, where all 23 respondents who completed a question on infrastructure challenges indicated one or more of these challenges. The finding that over a third of respondents do not submit their WGS sequence data to public repositories may be due to the reported internet connectivity challenges. The high demand for increased capacity in both WGS and bioinformatics analysis further underscores the broader infrastructure and skills gaps that may constrain the sustainability and expansion of genomic surveillance.

Logistical challenges were also noted, with reagent delivery times of more than 3 months and shelf-lives of 2–6 months commonly reported. It is therefore possible that many of these reagents may be expired or close to expiring on arrival, potentially disrupting laboratory workflows and hindering the continuity of genomic surveillance to influence healthcare or public health decisions. This highlights the need for reliable supply chains and reagents with longer shelf-lives to support sustainable genomic surveillance, which is a recognised challenge with healthcare diagnostics, vaccines and therapeutics more broadly on the continent.

Several limitations of the survey were identified, which restricted some of our analyses and may have influenced our findings. Due to the uneven spread of responses across the different African regions, including very few responses from Northern and Central Africa, we were unable to assess regional differences in laboratory capacity. This also means we may be misrepresenting capacity in Northern and Central African institutions. The purposive sampling approach and networks we had access to for distribution of the survey may have limited our ability to reach a wider range of laboratories.

Institutions with established microbiology services may have been more likely to engage with the survey, potentially resulting in an overestimation of blood culture and genomic capacity. Conversely, the survey did not assess whether laboratories referred specimens to external facilities for blood culture processing, which may have resulted in an underestimation of access to blood culture services. Furthermore, representing the amount of blood cultures testing positive as ranges of percentages rather than numbers may have been more suitable to enable a true estimation of blood culture positivity rates.

Respondents were able to select more than one laboratory type, since some institutions may comprise diagnostic, public health and research laboratory functions, even within the same department. However, questions relating to test capacity and equipment access did not require respondents to specify which laboratory had the reported capacity. Consequently, stratification by laboratory type for some of the results should be interpreted with caution. Similarly, the interpretation of the “internal” or “external” access options for WGS workflows and platforms may have varied between respondents, where a different laboratory in the same institution could have been perceived as internal by some respondents and external by others. At the question on the need for increased capacity for various steps of the WGS workflow, separating DNA extraction and library preparation may have been more appropriate. Capacity for DNA extraction may be more well-established and if external service providers are used for WGS, they would often accept DNA and perform the subsequent steps.

While this survey provides insight into the availability of microbiology and genomic capacity among African institutions, future studies should evaluate the quality of these services. Although referral pathways for genomic workflows were assessed, referral pathways for blood culture services and the frequency with which genomic workflows are used in routine practice warrant further investigation. Further investigation of supply chain challenges on the continent is needed, including establishing evidence-based shelf lives for commonly used laboratory consumables, as some reagents may still yield acceptable results for some time beyond their stated expiry dates.

## CONCLUSION

This survey demonstrated that capacity to support genomic surveillance of sepsis pathogens exists across participating African institutions, with more than half reporting access to blood culture and in-house genomic services. However, several challenges were identified that may limit the expansion and sustainability of genomic surveillance efforts.

Improving access to high-quality blood culture services is essential, not only for patient care but also for generating neonatal pathogen surveillance data. Beyond strengthening capacity to generate pathogen sequence data, efforts should focus on their timely analysis and dissemination to ensure translation into actionable insights that inform empiric neonatal sepsis treatment regimens and support neonatal unit outbreak investigations.

Standardised protocols for genomic surveillance of neonatal sepsis pathogens in Africa should prioritise widely accessible laboratory methods such as column-based DNA extraction, reagents with longer shelf-lives, and, where possible, bioinformatics workflows that minimise dependence on high-performance computing infrastructure or internet connectivity.

## Data Availability

The data underlying this research are available within the article and its Supporting Information files.

https://drive.google.com/file/d/1KT10y5GzU34edLXnHKY8Gc6urwIbPFyE/view?usp=share_link

https://drive.google.com/file/d/1VDE_UJ5JfH7Mfx32RN4KE5_CgQ3tD4IY/view?usp=share_link

## ACKNOWLEDGEMENTS

The authors would like to thank Dr Chiara Crestani (Institut Pasteur, Paris, France) and Dr Louise Teixeira Cerdeira (London School of Hygiene and Tropical Medicine, London, United Kingdom) for assistance with translation of the survey materials into French and Portuguese, respectively. The authors also appreciate the valuable input given by the Moleleki Manuscript Writing Group at the Division of Medical Microbiology and Immunology, Stellenbosch University.

## SUPPORTING INFORMATION

**S1 Appendix:** Informed consent and survey questions.

**S2 Appendix:** Supporting figures and table.

